# A Supervised Text Classification System Detects Fontan Patients in Electronic Records with Higher Accuracy than ICD Codes

**DOI:** 10.1101/2023.03.01.23286659

**Authors:** Y Guo, MA Al-Garadi, WM Book, LC Ivey, FH Rodriguez, CL Raskind-Hood, C Robichaux, A Sarker

**Affiliations:** Department of Biomedical Informatics, School of Medicine, Emory University, Atlanta, GA, United States; Vanderbilt University Medical Center, Vanderbilt University, Nashville, TN, United States; Department of Cardiology, School of Medicine, Emory University, Atlanta, GA, United States; Rollins School of Public Health, Emory University, Atlanta, GA, United States

**Keywords:** Natural language processing, Fontan, single ventricle, congenital heart disease

## Abstract

**Background:** The Fontan operation palliates single ventricle heart defects and is associated with significant morbidity and premature mortality. Native anatomy varies; thus, Fontan cases cannot always be identified by *International Classification of Diseases, Ninth and Tenth Revision, Clinical Modification (ICD-9-CM and ICD-10-CM)* codes, making it challenging to create large Fontan patient cohorts. We sought to develop natural language processing (NLP) based machine learning (ML) models, which utilize free text notes of patients, to automatically detect Fontan cases, and compare their performances with ICD code based classification.

**Methods and Results:** We included free text notes of 10,935 manually validated patients, of whom 778 (7.1%) were Fontan and 10,157 (92.9%) non-Fontan patients, from two large, diverse healthcare systems. Using 5-fold cross validation, we trained and evaluated multiple ML models, namely support vector machines (SVM) and a transformer based model for language understanding named RoBERTa (2 versions), for automatically identifying Fontan cases based on free text notes. To optimize classifier performances, we experimented with different text representation techniques, including a sliding window strategy to overcome the length limit imposed by RoBERTa. We compared the performances of the ML models to ICD code based classification using the F_1_ score metric. The ICD classification model, SVM, and RoBERTa achieved F_1_ scores of 0.81 (95% CI: 0.79-0.83), 0.95 (95% CI: 0.92-0.97), and 0.89 (95% CI: 0.88-0.85) for the positive (Fontan) class, respectively. SVM obtained the best performance (*p*<0.05), and both NLP models outperformed ICD code based classification (*p<0.05*). The novel sliding window strategy improved performance over the base RoBERTa model (*p<0.05*) but did not outperform SVM. ICD code based classification tended to have more false positives compared to both NLP models.

**Conclusions:** Our proposed NLP models can automatically detect Fontan patients based on clinical notes with higher accuracy than ICD codes. Since the sensitivity of ICD codes is high but the positive predictive value is low, it may be beneficial to apply ICD codes as a filter prior to applying NLP/ML to achieve optimal performance.

## Introduction

The Fontan operation is typically performed in early childhood to palliate single ventricle congenital heart defects (CHDs), with heterogeneous native anatomy. Following the Fontan operation, the absence of a pulsatile pulmonary circulation and chronically elevated hepatic venous pressures lead to numerous comorbid complications with age including arrhythmias, heart failure, hepatic fibrosis, renal disease, and other late complications^1^. The *International Classification of Diseases, Ninth and Tenth Revision, Clinical Modification (ICD-9-CM and ICD-10-CM)* diagnostic codes can be used to identify the categories of patients by their native anatomy and are commonly used in research for retrieving targeted cohort data. Additionally, ICD-9-CM and ICD-10-*Procedure Coding System* (PCS) procedural codes can be used to identify procedures a patient may have undergone. While all Fontan patients have similar post-surgical anatomy and complications regardless of the native anatomy^2^, not all patients with the same native anatomy require a Fontan operation. Therefore, it is difficult to identify Fontan cases strictly by their ICD codes.

Text notes associated with the electronic health records (EHRs) of Fontan patients typically contain lexical cues indicative of the Fontan operation, recognizable by subject matter experts. However, manually reviewing notes in EHRs is time-consuming, and the task is often impractical. Natural language processing (NLP) methods provide a potential solution to automatically detect evidence of Fontan for longitudinal cohort studies. NLP, a sub-discipline of computer science concerned with the use of computers to process information from human language, has been widely applied to clinical notes for creating healthcare applications. For example, studies have leveraged NLP of clinical notes for studying chronic diseases^3^, extracting critical human immunodeficiency virus and cardiovascular risk information^4,5^, analyzing critical limb ischemia^6^, and extracting ad-hoc concepts^7^. An effective NLP system, which can automatically identify Fontan cases from text notes in EHRs, will help improve the efficiency of creating Fontan cohorts, and hence, conducting health-related studies on Fontan cases.

In this paper, we sought to train and evaluate NLP based supervised ML systems to identify Fontan cases based on unstructured clinical notes in two large healthcare systems. We employed different text representation strategies and proposed a technological innovation to better model text using state-of-the-art transformer based text representation methods.

## Methods

### Machine Learning Classification Framework

The framework for the NLP based ML model training and evaluation is shown in Figure 1. The process includes three steps: (i) collecting clinical data, (ii) manually annotating whether a patient has had a Fontan operation or not, and (iii) training and evaluating supervised classification models. We provide further details in the following subsections.

**Figure 1:**
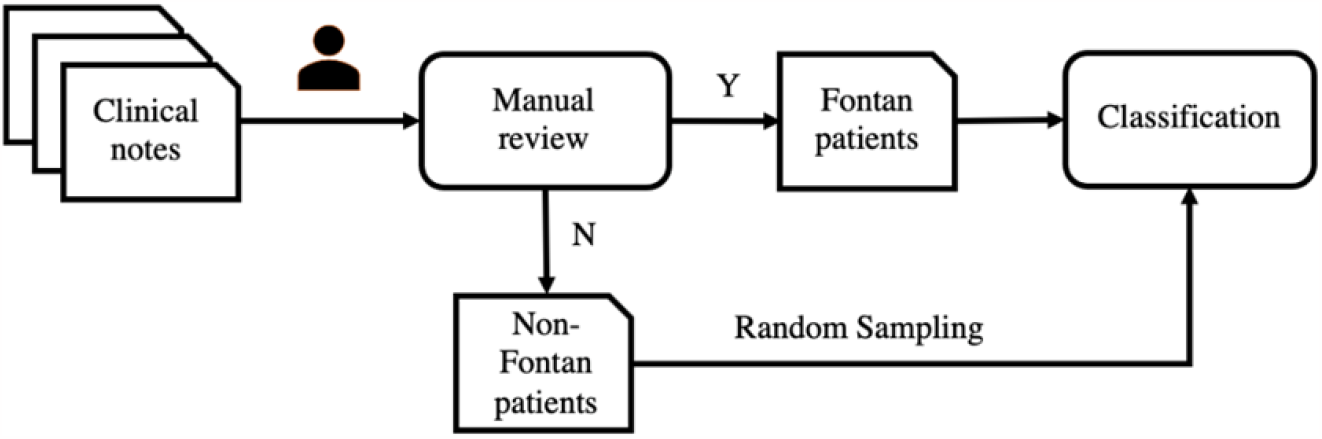
The development framework of the Fontan patient identification system.

### Study Population and Data Collection

This study utilized data on patients with at least one CHD ICD-9- or ICD-10-CM code (Table S1) documented in a healthcare encounter between 1/1/2010 and 12/31/2019 in databases for two state-wide multi-hospital tertiary healthcare systems, one pediatric (PHS) and one adult (AHS). Data were collected under a cooperative agreement with the Centers for Disease Control and Prevention (CDC-RFA-DD19-1902B). Emory University Institutional Review Board (IRB) approved the study on August 26, 2020 (IRB# STUDY00001030) and included a complete waiver of HIPAA authorization as well as waiver of informed consent. Manually abstracted Fontan cases were collected and managed using REDCap (Research Electronic Data Capture) at Emory University, a secure, web based software platform designed to support data capture for research studies.^8,9^

### Case Validation

Fontan cases were confirmed in one of four ways: (1) Their Fontan operation was identified and noted by clinicians and trained abstractors during a manual abstraction and clinical review of health records for 1500 CHD cases of all types in the PHS and AHS; (2) Their Fontan operation was documented in the pediatric STS database, which includes data, entered by trained research coordinators and clinicians and reviewed by a clinician, on diagnoses and surgeries of all pediatric congenital heart surgical cases in the United States; (3) The Fontan patient was included in an adult clinical Fontan tracking list created and maintained by clinicians to monitor clinical care for adult Fontan patients and verified by congenital cardiology clinicians; (4) An NLP-driven search was conducted on all text notes available for patients with at least one select single ventricle diagnosis code or Fontan procedure code (Table 1) who were not already labeled as Fontan cases from one of the first 3 mechanisms. The NLP-driven search performed *inexact* or *fuzzy matching* of the term ‘Fontan’ within the text of the notes so that words similar to Fontan, including potential misspellings, were captured. All retrieved posts were manually reviewed and Fontan cases were labeled. Reasons for false positives with NLP-driven search were noted, as were reasons for false negatives with ICD code based classification of single ventricle cases.

**Table 1:**
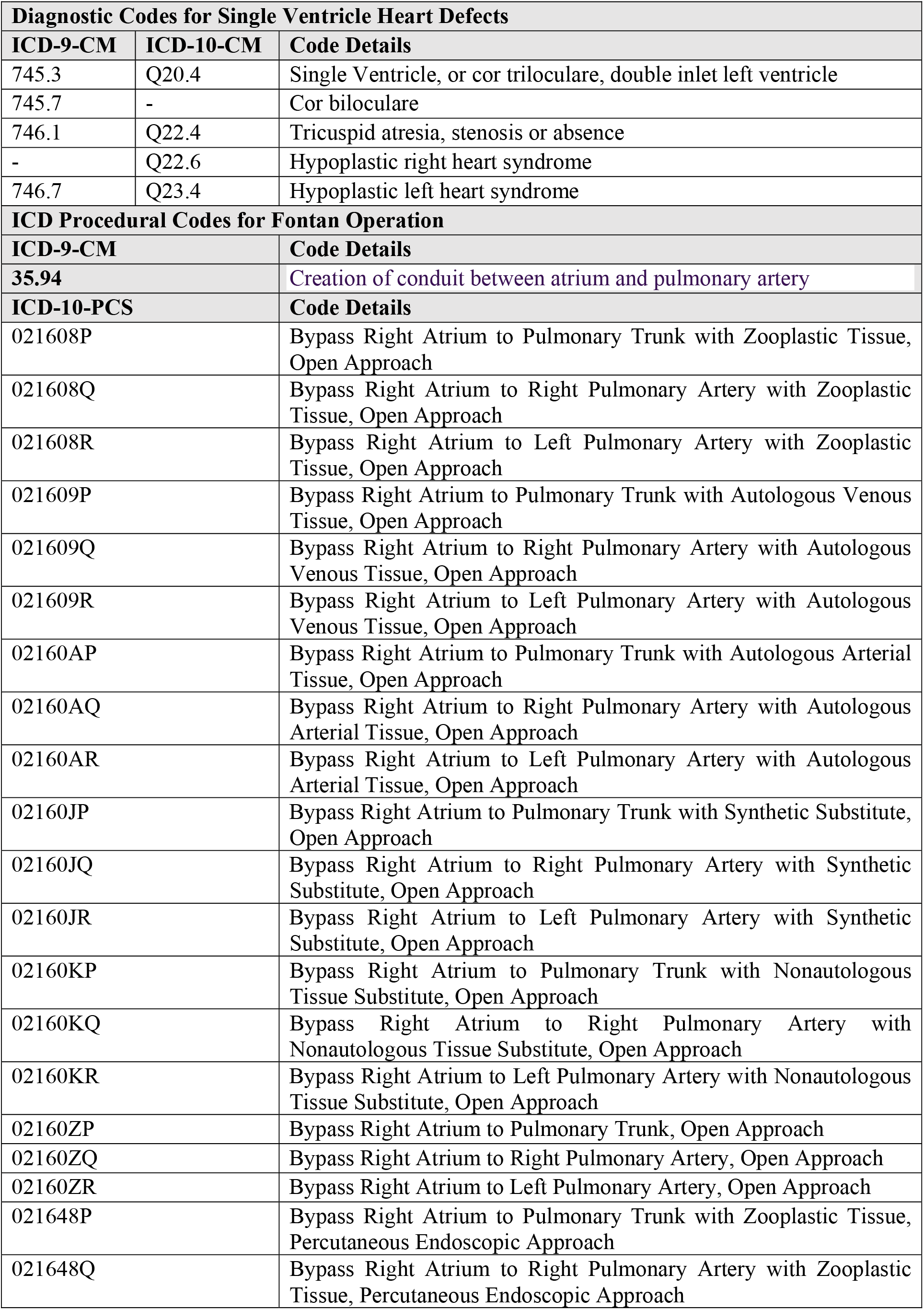
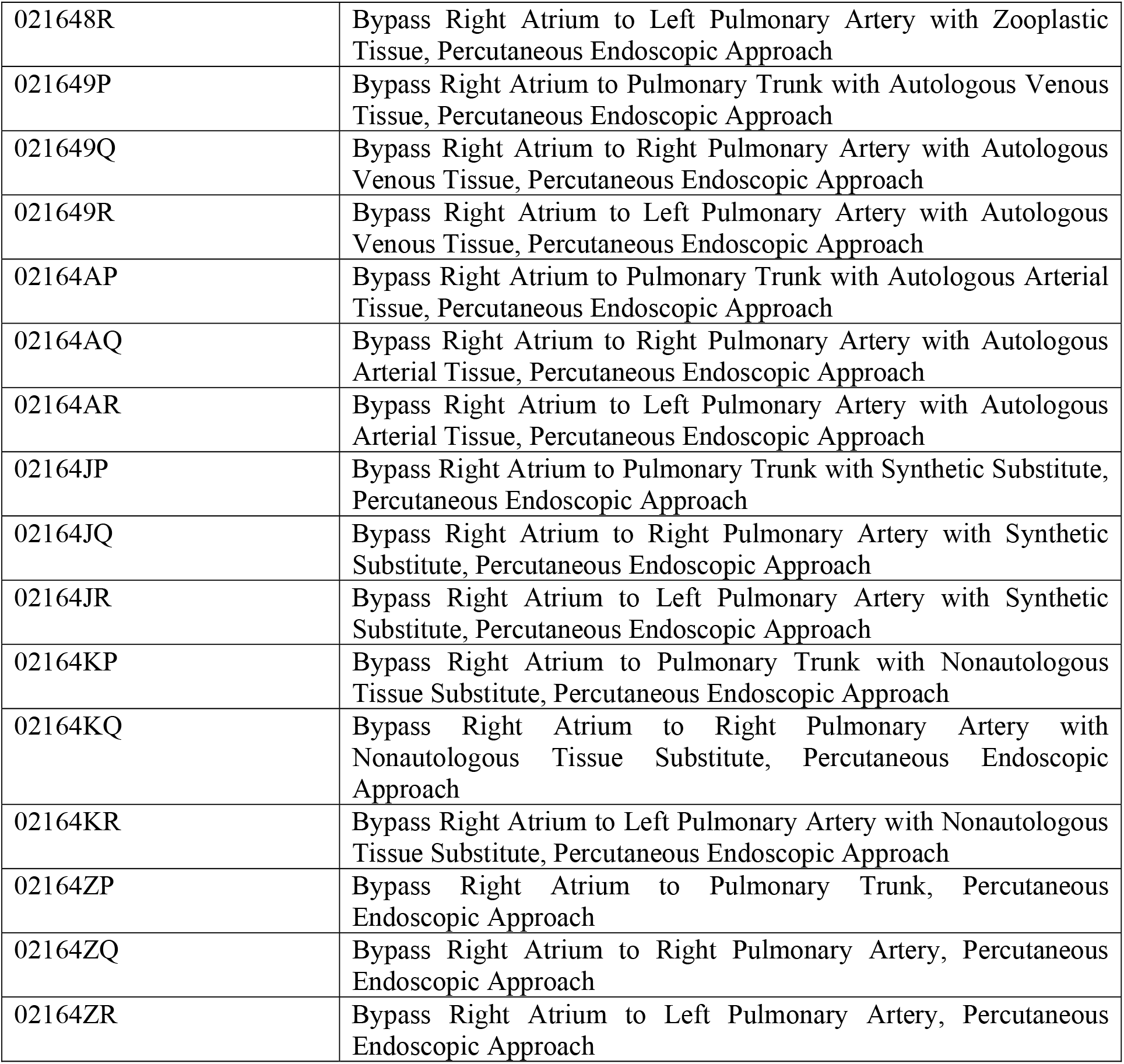
The ICD-9-CM and ICD-10-CM diagnostic codes and ICD-9 and ICD-10-PCS procedural codes used to classify Fontan cases for the ICD classifier?

| **Diagnostic Codes for Single Ventricle Heart Defects** |  |  |
| --- | --- | --- |
| **ICD-9-CM** | **ICD-10-CM** | **Code Details** |
| 745.3 | Q20.4 | Single Ventricle, or cor triloculare, double inlet left ventricle |
| 745.7 | - | Cor biloculare |
| 746.1 | Q22.4 | Tricuspid atresia, stenosis or absence |
| - | Q22.6 | Hypoplastic right heart syndrome |
| 746.7 | Q23.4 | Hypoplastic left heart syndrome |
| **ICD Procedural Codes for Fontan Operation** |  |  |
| **ICD-9-CM** |  | **Code Details** |
| **35.94** |  | Creation of conduit between atrium and pulmonary artery |
| **ICD-10-PCS** |  | **Code Details** |
| 021608P |  | Bypass Right Atrium to Pulmonary Trunk with Zooplastic Tissue, Open Approach |
| 021608Q |  | Bypass Right Atrium to Right Pulmonary Artery with Zooplastic Tissue, Open Approach |
| 021608R |  | Bypass Right Atrium to Left Pulmonary Artery with Zooplastic Tissue, Open Approach |
| 021609P |  | Bypass Right Atrium to Pulmonary Trunk with Autologous Venous Tissue, Open Approach |
| 021609Q |  | Bypass Right Atrium to Right Pulmonary Artery with Autologous Venous Tissue, Open Approach |
| 021609R |  | Bypass Right Atrium to Left Pulmonary Artery with Autologous Venous Tissue, Open Approach |
| 02160AP |  | Bypass Right Atrium to Pulmonary Trunk with Autologous Arterial Tissue, Open Approach |
| 02160AQ |  | Bypass Right Atrium to Right Pulmonary Artery with Autologous Arterial Tissue, Open Approach |
| 02160AR |  | Bypass Right Atrium to Left Pulmonary Artery with Autologous Arterial Tissue, Open Approach |
| 02160JP |  | Bypass Right Atrium to Pulmonary Trunk with Synthetic Substitute, Open Approach |
| 02160JQ |  | Bypass Right Atrium to Right Pulmonary Artery with Synthetic Substitute, Open Approach |
| 02160JR |  | Bypass Right Atrium to Left Pulmonary Artery with Synthetic Substitute, Open Approach |
| 02160KP |  | Bypass Right Atrium to Pulmonary Trunk with Nonautologous Tissue Substitute, Open Approach |
| 02160KQ |  | Bypass Right Atrium to Right Pulmonary Artery with Nonautologous Tissue Substitute, Open Approach |
| 02160KR |  | Bypass Right Atrium to Left Pulmonary Artery with Nonautologous Tissue Substitute, Open Approach |
| 02160ZP |  | Bypass Right Atrium to Pulmonary Trunk, Open Approach |
| 02160ZQ |  | Bypass Right Atrium to Right Pulmonary Artery, Open Approach |
| 02160ZR |  | Bypass Right Atrium to Left Pulmonary Artery, Open Approach |
| 021648P |  | Bypass Right Atrium to Pulmonary Trunk with Zooplastic Tissue, Percutaneous Endoscopic Approach |
| 021648Q |  | Bypass Right Atrium to Right Pulmonary Artery with Zooplastic Tissue, Percutaneous Endoscopic Approach |

|  |  |
| --- | --- |
| 021648R | Bypass Right Atrium to Left Pulmonary Artery with Zooplastic Tissue, Percutaneous Endoscopic Approach |
| 021649P | Bypass Right Atrium to Pulmonary Trunk with Autologous Venous Tissue, Percutaneous Endoscopic Approach |
| 021649Q | Bypass Right Atrium to Right Pulmonary Artery with Autologous Venous Tissue, Percutaneous Endoscopic Approach |
| 021649R | Bypass Right Atrium to Left Pulmonary Artery with Autologous Venous Tissue, Percutaneous Endoscopic Approach |
| 02164AP | Bypass Right Atrium to Pulmonary Trunk with Autologous Arterial Tissue, Percutaneous Endoscopic Approach |
| 02164AQ | Bypass Right Atrium to Right Pulmonary Artery with Autologous Arterial Tissue, Percutaneous Endoscopic Approach |
| 02164AR | Bypass Right Atrium to Left Pulmonary Artery with Autologous Arterial Tissue, Percutaneous Endoscopic Approach |
| 02164JP | Bypass Right Atrium to Pulmonary Trunk with Synthetic Substitute, Percutaneous Endoscopic Approach |
| 02164JQ | Bypass Right Atrium to Right Pulmonary Artery with Synthetic Substitute, Percutaneous Endoscopic Approach |
| 02164JR | Bypass Right Atrium to Left Pulmonary Artery with Synthetic Substitute, Percutaneous Endoscopic Approach |
| 02164KP | Bypass Right Atrium to Pulmonary Trunk with Nonautologous Tissue Substitute, Percutaneous Endoscopic Approach |
| 02164KQ | Bypass Right Atrium to Right Pulmonary Artery with Nonautologous Tissue Substitute, Percutaneous Endoscopic Approach |
| 02164KR | Bypass Right Atrium to Left Pulmonary Artery with Nonautologous Tissue Substitute, Percutaneous Endoscopic Approach |
| 02164ZP | Bypass Right Atrium to Pulmonary Trunk, Percutaneous Endoscopic Approach |
| 02164ZQ | Bypass Right Atrium to Right Pulmonary Artery, Percutaneous Endoscopic Approach |
| 02164ZR | Bypass Right Atrium to Left Pulmonary Artery, Percutaneous Endoscopic Approach |

### Classification Models

To automate the classification process and identify the best automatic classification strategy, we developed and evaluated multiple ML algorithms—Support Vector Machines (SVM) and a robustly optimized transformer based model for language understanding named RoBERTa (2 versions). Particularly, we compared what is an effective traditional text classification approach (SVM) to the latest and state-of-the-art approach (RoBERTa) currently known, and also compared the performance of the ML models to the ICD code based classification. For ML models, we used stratified 80-20 training-testing splits of data to ensure that the class distributions of the training and test sets remain the same. The model performances were measured by the precision (positive predictive value (PPV)), recall (sensitivity), and F_1_ score (harmonic mean of PPV and sensitivity) over the positive class on the test sets. We chose F_1_ score as the primary metric for comparison because it ensures that neither precision or recall is optimized at the expense of the other. Equations for computing these metrics are shown in Equation 1. For each classification model, we computed the performance on the test set for the cases from the AHS, the cases from the PHS, and the cases from both databases, respectively. Bootstrap resampling was used to compute 95% confidence intervals for the F_1_ scores.^10^ We performed significance testing for the F_1_ scores using the assumption-free randomization-based method proposed by Yeh (2000).^11,12^ The method computes a p-value based on the predictions of two models. If the p-value is less than 0.05, the difference in performance between the two models is significant. Further model-specific details are provided below.

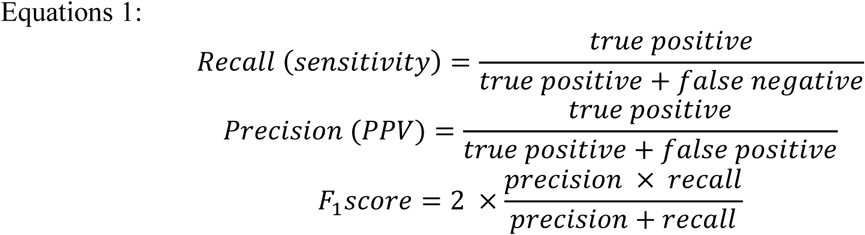

#### SVM Model

SVMs^13^ are popular choices when feature spaces are large, which is why they have been effective for text classification in the past and were chosen for this study. Term frequency–inverse document frequency (TF-IDF) for n-grams was used to obtain the vectorized representations of text notes used as features. An n-gram is a contiguous sequence of n words from the text. In our setting, we used 1-, 2-, 3-, and 4-grams. Term frequency– inverse document frequency is a numerical statistic used to measure the importance of a word in a document or set of documents. Term frequency (TF) refers to the number of times a word appears in a document (i.e., a clinical note). Inverse document frequency (IDF) refers to how rare the word is across the entire set of documents (i.e., our entire training dataset). TF-IDF is used to weigh the importance of words in a document, with respect to the entire set of documents. The TF-IDF vectors thus have higher numerical values for n-grams that are unique to each document and lower values for those occurring uniformly across all documents. During training, grid search was used to optimize two key hyper-parameters— kernel function *K* ∈ {linear, *RBF*} and *C* ∈ {2, 4, 6, 8}—and obtain the optimal setting for the model. Five-fold cross validation was used to evaluate the models by performing data-splitting 5 times, which can result in a less biased estimate of the model skill than using a static data split. For each data split, we trained the model on the training set, found the optimal model on the validation set, and evaluated the model on the testing set.

#### Transformer based Model

Transformer based models are relatively recent and have achieved state-of-the-art performances on many NLP tasks.^14–17^ As a representative of Transformer based models, we used RoBERTa^17^, a widely used pre-trained transformer based model, for Fontan classification. Instead of extracting features from text or generating n-grams, RoBERTa splits a clinical note into word pieces (i.e., tokens), encodes each token into a vector, and combines them into a vector representation for the clinical narrative, as shown in Figure 2. Unlike n-gram vectors, which are sparse in nature, the vectors generated by transformer based models are dense, and they also encode the context of each token (i.e., the same token will have a different representation based on the context in which it appears). A key limitation of the RoBERTa model is that it is limited to 512 tokens when representing texts in vector format. We found that most of the clinical notes exceeded that length limitation. This meant that using the standard RoBERTa model results in suboptimal or incomplete representations of the clinical notes. To overcome this limitation, we used a sliding window strategy to split the long notes into multiple partial notes. The sliding window was of size 512 tokens, and when passed through the entire note, each unique 512 token sequence in it was represented as an individual document. Thus, the model could be applied independently to each of the subdocuments represented by the window, and it could make independent predictions for each. In our approach, after the model predicted labels for each subdocument independently, the mode of the predictions over all the sub-sequences as the final prediction (i.e., majority voting) was used. Like SVM, 5-fold cross validation was used to evaluate the model. For each data split, we trained the model for 5 rounds. For each round, the model went through the training data once. We found the optimal model checkpoint on the validation set and evaluated the model on the test set. Other hyper-parameters and technical details are presented in Table S2 of the supplementary material.

**Figure 2:**
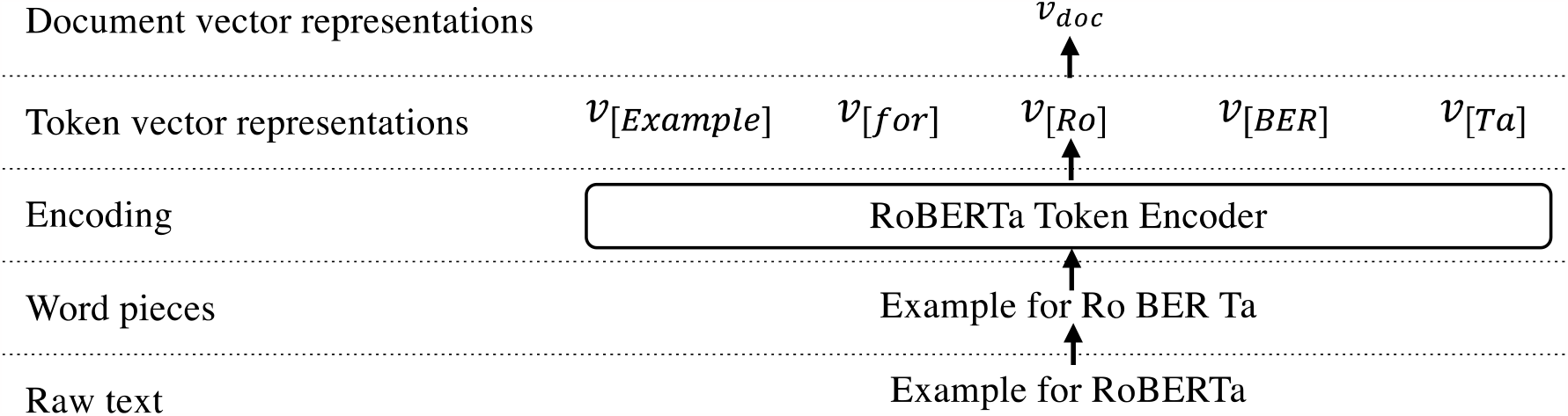
The process of how RoBERTa converts a clinical narrative into a vector representation.

#### ICD Code based Classification Model

We developed a classification model that used only the ICD codes in Table 1 associated with each health record to identify potential Fontan patients. ICD-9-CM and ICD-10-CM diagnostic codes for hypoplastic left heart syndrome (746.7/Q23.4), tricuspid atresia (746.1/Q22.4/Q22.6), and double inlet left ventricle/single ventricle (745.3/Q20.4) were included as individuals with these single ventricle heart defects typically undergo Fontan palliation in childhood. ICD-9-CM and ICD-10-PCS procedural codes for the Fontan operation in Table 1 were also used to identify Fontan cases based on the presence of a Fontan operation procedural code associated with the case.^18^ Depending on the severity of their specific heart defect, individuals with double outlet right ventricle, atrioventricular canal defect, pulmonary atresia intact ventricular septum, and other native anatomy may undergo Fontan palliation or two-ventricle repair; However ICD codes do not capture severity of any given anatomic defect to differentiate those who would receive Fontan from two ventricle repair so codes for these defects were not used to identify Fontan cases in this classification scheme. If any code associated with a patient’s record had at least one of the specific single ventricle codes included in Table 1 and/or the ICD-10-PCS Fontan operation codes noted in Table 1, the patient was classified as a Fontan case for this model. Different from the ML models, the ICD code classification was unsupervised, and there was no need to perform training-testing data splitting. Therefore, we evaluated the ICD code based classification model on the whole dataset.

### Post-classification Analyses

#### Learning Curve Analysis

To the best of our knowledge, no prior research effort has attempted to automate the task of building a Fontan cohort via supervised ML using the text data in their EHRs. Consequently, we had to annotate our own data. When annotating a sample of texts for a task like this, an important question arises about the amount of data needed to annotate. More specifically, at what number of annotations does the ML model start plateauing? To investigate this, the model performances were evaluated at different sizes of training data (20%, 40%, 60%, 80%, 100%) while keeping the test set identical. We anticipated that this analysis may provide some insights about whether further increasing the training data size is necessary to improve the models’ performances, or at what data size the model starts performing with sufficient accuracy.

#### Error Analysis

We manually analyzed the patient records that were misclassified by the ML models and summarized the common patterns. The differences between the characteristics of the clinical notes from the AHS and the PHS were also analyzed, since these could have affected the model performance. This analysis was performed particularly to identify the limitations of the model and any potential bias. Knowledge about the causes of errors may guide future directions of research to improve the model.

## Results

A total of 10,935 cases with available text notes, 778 validated Fontan and 10,157 non-Fontan, were identified following de-duplication and linked back to encounter-level data and text notes from the EHR. Of these cases, 210 Fontan and 7400 non-Fontan cases were from the AHS, and 568 Fontan and 2757 non-Fontan cases were from the PHS. Cases that could not be linked back to text notes or without ICD codes available were excluded (n=25,066) from the dataset for the NLP system development and evaluation. The average length of clinical notes from the AHS was 35 words, while that from the PHS was 1107 words.

### Classification Results

The classification results for the ML models and the ICD code based classification model are shown in Table 2, including the precision, recall, F_1_ score, and 95% confidence interval for the F_1_ score for each model. The exact p-values of the significance tests for the difference in performance between pairs of models are shown in Table 3. For both AHS and PHS data combined, the ICD code classification model had a precision score of 0.74, a recall score of 0.90, and a F_1_ score 0.81 (95% CI: 0.79-0.83). The ML models significantly outperformed the model using ICD codes (*p < 0*.*05*). The model performance of RoBERTa with sliding window was better than that without sliding window (*p < 0*.*05*), especially for the recall score (0.81 vs 0.51 in AHS and PHS combined). The results show that applying the sliding window strategy to RoBERTa can overcome the limitation of the default model and particularly improve the model’s ability to find true positives. SVM significantly outperformed RoBERTa (*p<0*.*05*), demonstrating that the traditional model is better able to capture the meanings of the texts compared to the more recently proposed transformer based RoBERTa model that uses a pretrained language model. For both AHS and PHS data combined, precision for SVM was 0.97, recall was 0.95, and F_1_ score was 0.95 (95% CI:0.92-0.97) (Table 2). The confusion matrices for SVM and RoBERTa are shown in Figure 3 to visualize the high accuracy of SVM as the best-performing model and the performance gap between SVM and RoBERTa.

**Table 2:**
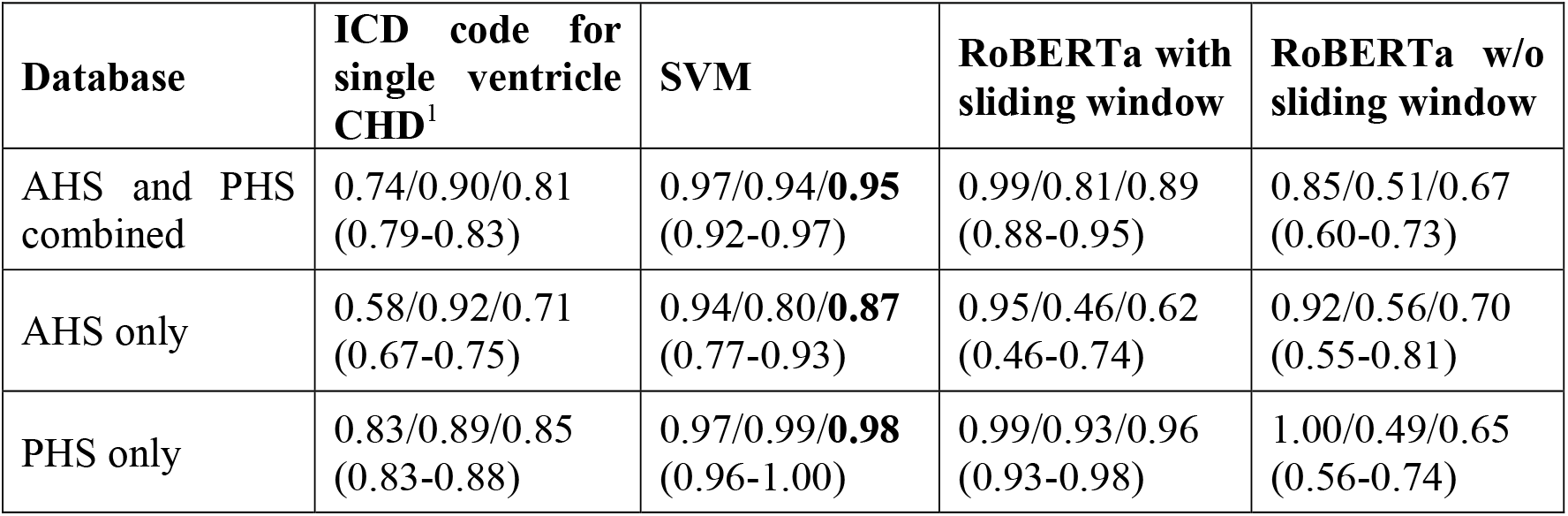
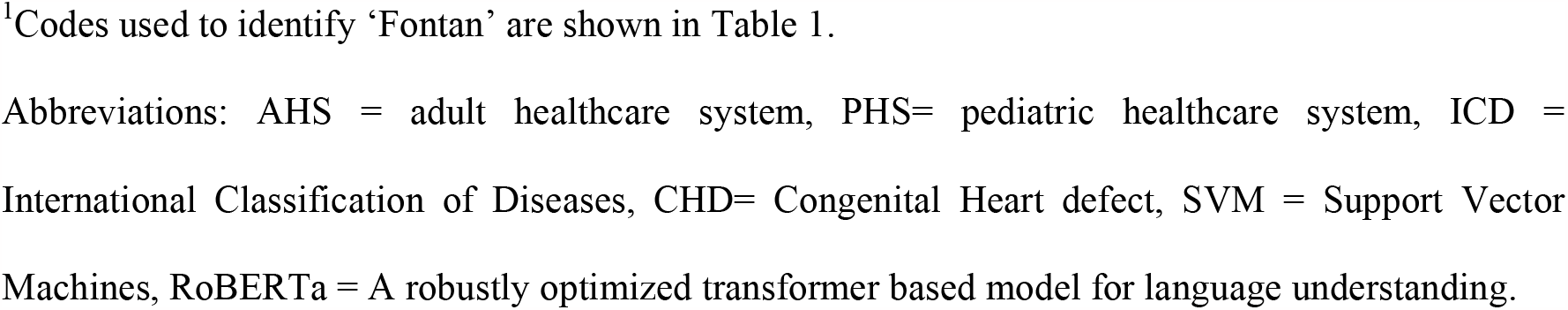
The results of ICD code classifiers and ML models. The model performances were presented as *precision/recall/F*_*1*_ *score (95% bootstrap confidence interval)*. The best model performance for each database is in bold. ^1^Codes used to identify ‘Fontan’ are shown in Table 1. Abbreviations: AHS = adult healthcare system, PHS= pediatric healthcare system, ICD = International Classification of Diseases, CHD= Congenital Heart defect, SVM = Support Vector Machines, RoBERTa = A robustly optimized transformer based model for language understanding.

**Table 3:** The significance testing for the difference in performance between two models.

| Model 1 | Model 2 | p-value |
| --- | --- | --- |
| SVM | RoBERTa with sliding window | 0.0003 |
| SVM | RoBERTa w/o sliding window | 0.0001 |
| SVM | ICD code for single ventricle CHD | 0.0001 |
| RoBERTa with sliding window | RoBERTa w/o sliding window | 0.0001 |
| RoBERTa with sliding window | ICD code for single ventricle CHD | 0.0001 |
| RoBERTa w/o sliding window | ICD code for single ventricle CHD | 0.0001 |

**Figure 3:**
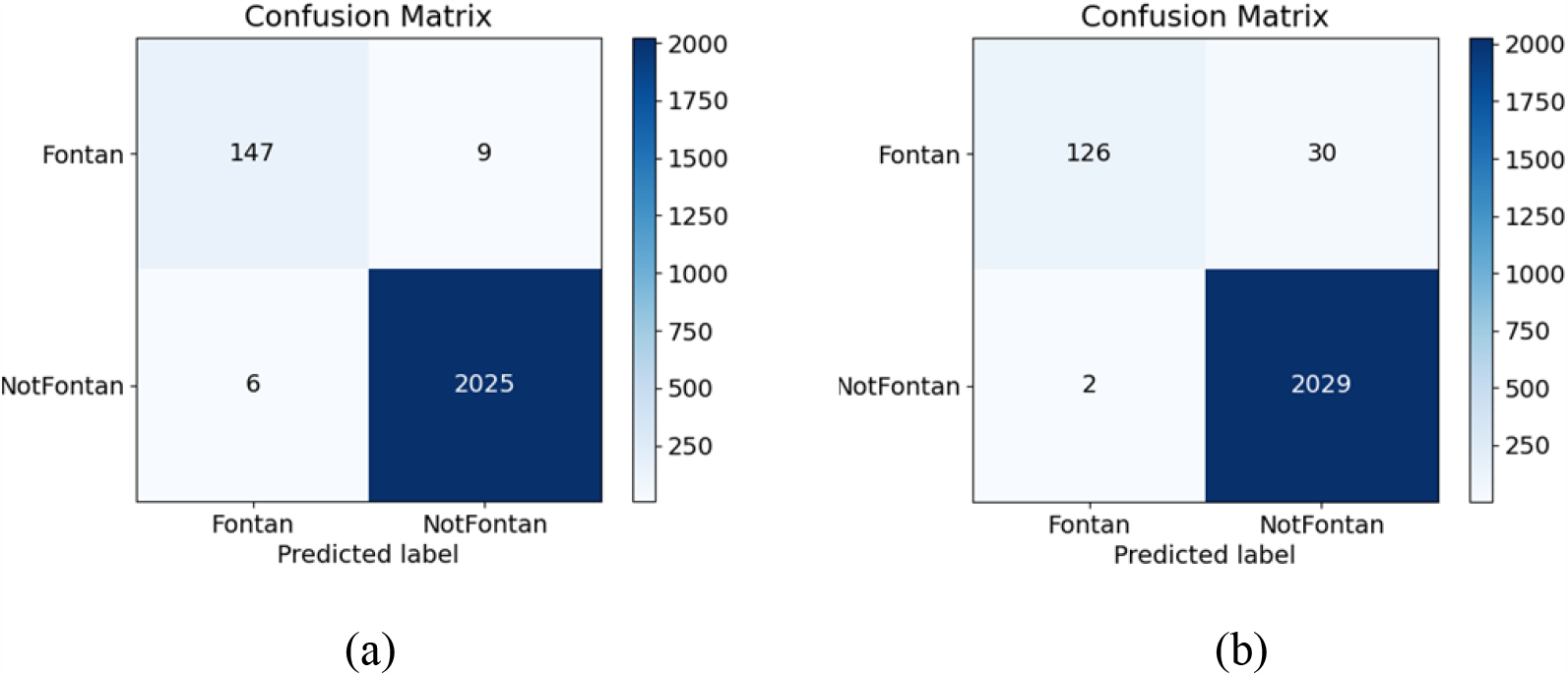
The confusion matrices curve of SVM (a) and RoBERTa (b).

### Post-classification Analysis

#### Error Analysis of Machine Learning Models

The confusion matrices reveal that SVM predicted more true positives and fewer false negatives than RoBERTa. This may mean that SVM is better at detecting non-Fontan cases compared to RoBERTa. RoBERTa with sliding window significantly underperformed compared to SVM on the cases from the AHS but achieved comparable results on the cases from the PHS. In contrast, RoBERTa without sliding window significantly underperformed compared to SVM on the cases from both sites. The performance gap can be attributed to the difference between the average lengths of the clinical notes from the two databases—the clinical notes from the PHS were substantially longer than those from the AHS in our dataset. The results suggest that although applying the sliding window strategy can help the model performance for long texts, it might lead to suboptimal representation for short texts. Further work is required to explore how to address the challenge of long clinical notes.

### Learning Curve

For RoBERTa (with sliding window) and SVM, we evaluated the model performances at different sizes of training data (20%, 40%, 60%, 80%, 100%) on the same test set for which performance is reported in Table 2. Figure 4 presents the F_1_ scores on the test set for different training set sizes. SVM consistently performed better than the RoBERTa model although the latter showed greater improvement as more training data was added. This suggests that while the SVM classifier performs better with the dataset currently available, it is possible that with more training data, the RoBERTa model may outperform it.

**Figure 4:**
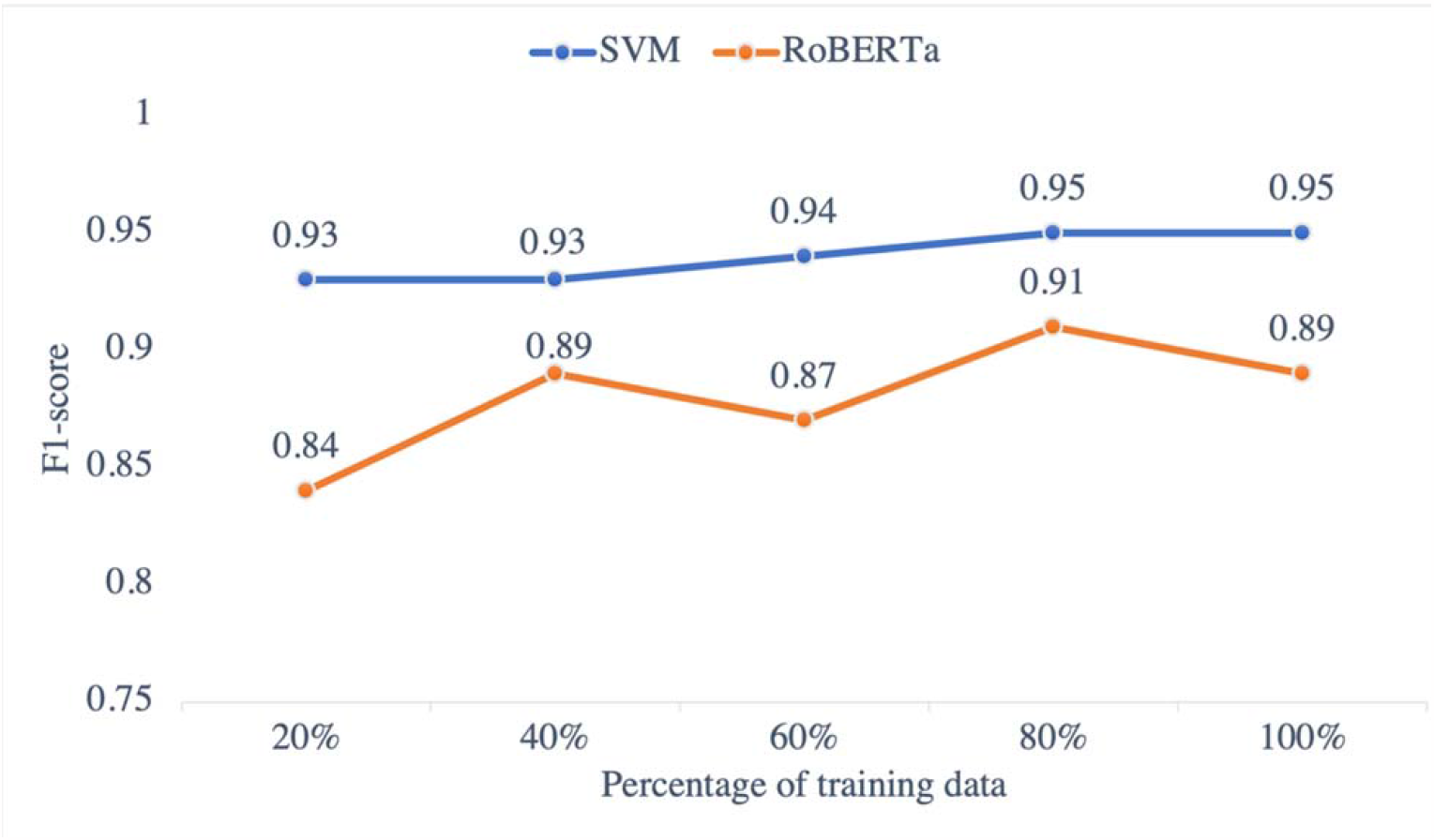
The model performance of SVM and RoBERTa using different percentage of training data.

## Discussion

Our experiments demonstrate that NLP based ML is a feasible, effective, and accurate mechanism for detecting Fontan cases based on the text notes of the EHRs. When analyzing the cases which were predicted as negative by the ICD code classifier but positive by the best-performing SVM classifier, we found that most of these cases were true positives, illustrating that SVM is better at identifying Fontan cases than an ICD code based classification. The high classification F_1_ score of 0.95 suggests this is a reliable method to accurately classify Fontan cases in EHRs based on text notes. The learning curve analysis showed that despite the strong performance, there maybe room for further improvement, particularly for transformer based models.

The ICD code based classification had several limitations leading to both false positives and false negatives. When using a combination of ICD-9-CM and ICD-10-CM/PCS diagnostic and procedural codes to identify Fontan cases, the PPV of ICD codes to detect Fontan cases was 58% in the AHS dataset, 83% in the PHS dataset, and 74% in the combined dataset. ICD diagnostic codes classify cases by native anatomy, which can vary in severity not accounted for by ICD codes. Thus, while some patients may require a Fontan operation, others may not despite having the same anatomic CHD. While there are ICD procedural codes for the Fontan operation,^18^ these are typically only used at the time of the operation, limiting their usefulness for cases when the operation occurred outside of the surveillance period or outside of the dataset. If the operation did not occur within the surveillance window and dataset, the only code based indicators that may be used in follow-up encounters to document prior Fontan operation are non-specific diagnosis codes for “*Personal history of congenital heart surgery*” (i.e., V13.65 and Z87.74). Researchers may instead decide to identify Fontan patients using only ICD codes for hypoplastic left heart syndrome, tricuspid atresia, double inlet left ventricle and single ventricle, since patients with these defects most likely have had a Fontan operation; however, individuals with other heart defects that may undergo Fontan operation, such as unbalanced atrioventricular canal defect or pulmonary atresia with intact ventricular septum, would be missed. Alternatively, including more ICD codes for these other heart defects to identify Fontan cases would result in false positives, since some of these cases may have two ventricle repair in place of Fontan palliation depending on the severity of their specific heart defect. Understanding longitudinal and late outcomes of Fontan patients with administrative data is limited by the poor PPV of ICD codes to identify the cohort.

Applying NLP based ML models on the clinical notes can improve the precision of the system, reduce the risk of mistakenly detecting non-Fontan patients, and capture Fontan cases missed by an ICD code based approach. This work is an example of how NLP is more accurate than the ICD code based approach for identifying individuals with single ventricle heart defects who have undergone Fontan palliation, allowing identification of more cases with greater accuracy. Individuals who have undergone Fontan palliation face complications that increase with time following Fontan operations.^19^ Limited data is available to understand factors that contribute to these adverse outcomes. The NLP-driven methodology that we have developed may improve longitudinal surveillance of individuals who have undergone Fontan palliation in childhood, allowing the incorporation of individuals who may be followed outside of tertiary referral centers.

### Strengths and Limitations

The primary strengths of our study are summarized below:

- We are the first study to collect clinical notes, develop annotation guidelines, and create an annotated dataset to develop NLP based supervised classification models for Fontan case detection.
- We developed and evaluated two supervised classification models—SVM and RoBERTa, which can more effectively detect Fontan patients based on clinical notes with high F_1_ score compared to ICD codes.
- We proposed a strategy for improving the modeling of long texts by the RoBERTa classifier. The strategy uses a windowing method followed by majority voting to optimize the performance of the classifier.
- We experimented with data from two diverse health systems and our best-performing model demonstrated relatively low variance across the datasets, suggesting that the NLP model is generalizable across sites.

The applicability of our study is limited by the requirement of availability of text notes in EHRs. Cases without available text notes could not be included in our cohort. Possible reasons for missing text notes include an earlier era of medical record systems which may have contained scanned notes, or legacy medical records systems that lack access to older notes. However, as almost all health systems are electronic in today’s world, NLP based methods for cohort creation will have high utility beyond the scope of this study. Although we are unable to share the data for this study, our code has been made publicly available, and researchers with annotated Fontan data from other health systems may adapt our code for their studies with relative ease.

NLP could be used as an adjunct to detect Fontan cases in a cohort initially identified by CHD ICD codes noted in Table S1. While the accuracy of the NLP/ML model is close to perfect, it is not correct 100% of the time, so some false positives/negatives are possible. It is also possible that the performance of the trained NLP model may be lower when applied to data from locations other than the two included in this study. This is a relatively common scenario for NLP models when they are trained at one site and evaluated at a different one. In this study, the evaluation on data from two vastly different health systems illustrates that the automated method is portable, although future research is necessary to address some of the limitations of the state-of-the-art NLP models, such as the length limitation.

## Conclusions

We developed and evaluated two NLP based ML models—SVM and RoBERTa, which can more effectively detect Fontan patients based on clinical notes with higher accuracy than ICD codes. Our experiments suggested that since the sensitivity of ICD codes is high but PPV is low, it may be beneficial to apply ICD codes as a filter prior to applying NLP/ML to improve performance. Accurate identification of a Fontan cohort enables the development of large Fontan datasets to understand longitudinal outcomes in this population. Our code is available at https://github.com/yguo0102/Fontan_classification.

## Data Availability

As per Emory University policy, patient data will not be shared with researchers outside the Emory Health system. Collaborators interested in joining the studies will be reviewed on a case by case basis and will only be allowed to access relevant data following approval from Emory University IRB. All code associated with the natural language processing and machine learning experiments is publicly available (see GitHub link in paper).

https://github.com/yguo0102/Fontan_classification

## Acknowledgments

TBA

